# Exposome-wide gene-environment interaction study of psychotic experiences in the UK Biobank

**DOI:** 10.1101/2024.09.06.24313177

**Authors:** Bochao Danae Lin, Lotta-Katrin Pries, Angelo Arias-Magnasco, Boris Klingenberg, David E. J. Linden, Gabriella A. M. Blokland, Dennis van der Meer, Jurjen J. Luykx, Bart P. F. Rutten, Sinan Guloksuz

## Abstract

**Background:** A previous study successfully identified 148 out of 23,098 exposures associated with any psychotic experiences (PE) in the UK Biobank using an exposome-wide association study (XWAS). Research has shown that the polygenic risk score for schizophrenia (PRS-SCZ) is associated with PE. However, the interaction of these exposures and PRS-SCZ remains unknown.

**Method:** To systematically investigate gene-environment interaction underlying PE through data-driven agnostic analyses, we conducted 1) a conditional XWAS adjusting for PRS-SCZ to estimate the main effects of the exposures and PRS-SCZ, respectively; 2) exposome-wide interaction studies (XWIS) to estimate multiplicative and additive interactions between PRS-SCZ and exposures; and 3) the correlations between PRS-SCZ and exposures. The study included 148,502 participants from UK biobank.

**Results:** In the conditional XWAS models, the significant effects of PRS-SCZ and 148 exposures on PE remained statistically significant. In the XWIS model, we found a significant multiplicative (Ms, 1.23, 95%CI, 1.10-1.37; P=4.0×10^-4^) and additive (RERI, 0.55; 95%CI, 0.32-0.77; SI, 0.22; 95%CI, 0.14-0.30; AP, 1.59; 95%CI, 1.30-1.91; all P < 0.05/148) interaction between PRS-SCZ and variable “serious medical conditions or disability” on PE. There were six additive gene-environment interactions identified for mental distress, help/treatment-seeking behaviors, vitamin D and sleep problems. In the correlation test focused on seven exposures with significant interaction with PRS-SCZ, no significant or small (r2< 0.04) gene-environment correlations were estimated.

**Conclusion:** These findings reveal preliminary evidence for gene-environment interaction underlying PEs and suggest that genetic vulnerability and exposures might represent intertwined pathways leading to psychosis.

## Introduction

Psychotic experiences (PE), characterized by delusions (unreal beliefs or impressions) or hallucinations (unreal visual or auditory perceptions), are common and disabling conditions with a prevalence of 5-10% in the general population (1). Behavioral, genetic and epidemiological research found that PE might represent subtle, subclinical symptoms across the psychosis spectrum and often precede or accompany the onset of clinical psychosis(2). Longitudinal studies and familial aggregation research suggest a substantial overlap between PE and the development of schizophrenia spectrum disorders(3, 4). PEs are moderately heritable and show considerable environmental influence (5) with well-established heritability and environmental risk factors. Understanding the underlying mechanisms and genetic and environmental correlates of psychotic experiences is crucial for the development of tailored prevention, targeted interventions and the enhancement of clinical outcomes in individuals with mental health disorders.

Hypothesis-driven research has identified several environmental factors associated with psychosis such as bullying (6), stressful life events (7), cannabis use (8), tobacco use (9), and low birth weight (10), as well as less studied exposures such as physical activity (11), toxins (12), and nutrients. However, these one-exposure-to-one-outcome hypothesis-testing studies fail to embrace the multiplicity of exposures and are prone to selective reporting and publication bias, involving arbitrary decisions. The availability of large public datasets, along with increased transparency in data processing and standardized analytical algorithms of agnostic data-driven approaches have increased reproducibility. A recent exposome-wide analysis of PE in the UK-Biobank has confirmed previous environmental factors associated with PE, as well as factors that have not been considered thus far, such as major dietary changes in the last 5 years, and playing computer games (13).

The relative contribution of genetic influences to psychotic experiences was considered relatively small with a SNP heritability < 2% (14, 15). Although the GWAS of PE identified genome-wide significant loci, none showed evidence of colocalization with schizophrenia (16). Significant genetic correlations (rg) between any PE and psychiatric diseases such as schizophrenia have been detected. However, findings of studies investigating the associations between PE and polygenic risk score for schizophrenia (PRS-SCZ) have been inconsistent, showing no (17, 18), weak (15, 16), or significant positive association(19, 20). These studies might be limited by the statistical power of GWAS and target sample size. Therefore, the association of PRS-SCZ with PE remains to be verified using the most recent SCZ GWAS(21).

In a twin study of PE (20), heritability decreased with increasing environmental exposure, highlighting the importance of a diathesis-stress or bioecological framework for understanding adolescent PE. Previous candidate gene-environment interactions (GxE) studies of PE have yielded inconsistent results (22–25). The advent of polygenic scores (PRS), which aggregate genome-wide common variants to index a person’s genetic propensities for a trait, has created opportunities for testing GxE. Recent GxE studies testing the interaction of PRS-SCZ with high birth weight (26) and smoking (27) also need to be replicated. Hereby, we conducted the first systematic and agnostic exposome-wide interaction analyses to identify the gene-environment interaction underlying PE.

## Methods and Materials

### Sample

The current study included participants from the UK Biobank (UKB), a large prospective population-based cohort that included around half a million participants from the United Kingdom (28). All participants provided written consent and ethical approval was given by the National Research Ethics Service Committee North West Multi-Centre Haydock, Committee reference: 11/NW/0382 (29). The current study (UKB project number: 55392) analyzed participants with complete data on the Mental Health Questionnaire (29) that assessed PE (n=155,247; 57% female; mean age=55.94[SD=7.74] years).

### Psychotic experiences

Guided by previous reports (13, 16, 27), a binary variable of any PE (n=7,803) was defined as an endorsement of any of the following four-lifetime items: visual hallucination, auditory hallucination, reference delusion, and persecutory delusion, variables namely, ever seen an un-real vision, ever heard an un-real voice, ever believed in un-real communications or signs, and ever believed in an un-real conspiracy against self.

### Correlates of psychotic experiences

For the current analyses, we included 148 variables (**supplementary material and eTable 1**) which were significantly associated with PE in the previous exposome-wide association study (XWAS) after applying Bonferroni correction (13). These 148 exposures, consisting of 109 binary and 39 continuous variables, belong to 13 UKB categories including environmental, lifestyle, behavioral, and sociodemographic factors. Most exposures are associated with increased PE, except for 26 exposures (such as vitamin D intake, and general health rating) that were associated with decreased PE.

In this study, we further dichotomized the 39 continuous exposures at the 75th percentile assigning values of 1 and 0, concordant with our previous GxE analyses (30, 31). As previously suggested for additive interactions, we reverse-coded the 26 negative correlates of PE, with 1 indicating “high-risk” and 0 indicating “low-risk” (32). This approach was used across all analyses to ensure comparable and consistent results (32) Therefore, the direction of effects of these 26 correlates on PE differs from the previous study (**eTable 2**).

### Polygenic risk score estimation

We calculated the PRS-SCZ for 151,627 participants who had available genetic and phenotypic information. We used summary statistics from recent GWAS of schizophrenia derived from European-ancestry (21) to calculate PRS-SCZ. To estimate the PRSs, we used PRS-continuous shrinkage (33) (PRS-cs-auto) as the main analyses and PRSice2 (p-value threshold =0.05) (34) as sensitivity analyses. Similar to previous studies, PRS-SCZs were dichotomized at the 75th percentile (30, 31) (hereafter PRS-cs-auto-SCZ_75_ and PRSice-SCZ_75_). Detailed methodology can be found in the supplementary material.

### Statistical analyses

Analyses were performed using R (version 4.0.4) (35) from November 1, 2023, to February 1, 2024. There were three sequential analytical steps (**eFigure 1**). First, we tested the main effects of PRS-SCZ on PE using baseline logistic models with covariates, including sex, age and first 3 genetic Principal components (PCs) (PE ∼ sex + age + PC1 + PC2 + PC3 + PRS-SCZ). Second, we added each of the 148 exposures into PRS (PE∼ sex + age + PC1 + PC2 + PC3 + PRS-SCZ + exposure)., using the "interactionR” (36) to estimate GxE interactions. Third, the correlations of PRS-SCZ with each of the 148 exposures were estimated using Pearson correlation. Bonferroni correction was applied for adjusting multiple testing (p <0.05/148). We also attempted to replicate previously demonstrated gene-environment interactions: birth weight and smoking behavior (26, 27). Sensitivity analyses were conducted using PRSice-SCZ_75_ across all analytical steps.

### Multiplicative and additive interaction

Interactions on the multiplicative scale assess whether the joint effect of the PRS and exposure is greater than the product of their individual effects. For multiplicative interaction, we integrated a product term (Multiplicative scale, Ms) of PRS-SCZ with each exposure on PE in the logistic regression models. Besides the Ms coefficients, corresponding p-values and 95% confidence intervals (CIs) were also reported.

Interactions on the additive scale assess whether the joint effect of exposure and the PRS-SCZ is greater than the sum of their individual effects. Relative excess risk due to interaction (RERI), attributable proportion of interaction (AP), and synergy index (SI), along with corresponding p-values and 95% CIs were utilized to perform effect modification analysis on the additive scale. We also estimated ORs, 95% CIs, and P values in each exposure and PRS strata to evaluate whether the effect of the exposure differed within the strata of PRS-SCZ. To estimate the CIs for the additive interactions, the simple asymptotic delta method (37) and the variance recovery (‘MOVER’) method (38) were applied. As sensitivity analyses, we additionally estimated the CIs of the interactions with the 39 continuous exposures and the continuous PRSs using the non-parametric bootstrapping method with 1000 bootstrap resampling (39).

### Replication of previous gene-environment interactions

Recent studies have shown GxE in PE related to birth weight and smoking behavior, employing PRS-SCZ derived from PGC2 (40) across different datasets. In addition to the exposome-wide interaction analyses, we attempted to replicate the findings using PRS-SCZ PGC3 (21) to confirm previous findings.

## Results

### Main effects

In the baseline model, PRS-cs-auto-SCZ_75_ significantly predicted PE (OR, 1.14; 95 %CI, 1.11-1.17; P=7.27×10^-24^, R^2^=0.21%). In the 148 conditional XWAS models, adding exposure into the logistic models, the significant effects of PRS-cs-auto-SCZ_75_ on PE remained significant (ORs, 1.11-1.15; R^2^, 0.12-0.24%; P-values 6.6 ×10^-13^ to 1.1×10^-26^; **eTable 3**). Under the condition of PRS-cs-auto-SCZ_75_, all the ORs of the 148 exposures remained significant (**eFigure 2**). The sensitivity analyses with PRSice-SCZ_75_ confirmed these results. **eTable 3**).

### Multiplicative scales

Among the 148 exposures, the only significant multiplicative interaction with PRS-SCZ was found for disability (“other serious medical condition disability diagnosed by doctor with the Multiplicative scale (Ms) 1.23 (**Table 1**, 95 %CI, 1.10-1.37; P=4.0×10^-4^). Four analyses indicated nominally statistically significant interactions **(Table 2)** for visiting a psychiatrist for mental health, mental distress, vitamin D, and visiting a GP for mental health. In the sensitivity test using PRSice-SCZ_75_, the top multiplicative interaction remained disability (Ms, 1,22; 95 %CI, 1,092-1,37, P=0.0005), but it was not statistically significant after Bonferroni correction. Furthermore, three nominal significant interactions remained: mental distress, visiting a psychiatrist for mental health, and vitamin D (**eTable 4**).

**Table 1.** Interaction of disability and PRS-cs-auto-SCZ_75_ on psychotic experience.

|  | Disability: No | Disability: Yes | Disability Yes, vs No within strata of PRSice-SCZ |
| --- | --- | --- | --- |
|  | OR [95%CI]<br>P | OR [95%CI]<br>P | OR [95%CI]<br>P |
| PRS-cs-auto-SCZ<sub>75</sub> =0 | 1[Reference] | 1.82[1.72-1.94]<br>P<10<sup>-5</sup> | 1.82[1.74-1.94]<br>P<10<sup>-5</sup> |
| PRS-cs-auto-SCZ<sub>75</sub> =1 | 1.10[1.04-1.17]<br>P=0.023 | 2.47[2.27-2.69]<br>P<10<sup>-5</sup> | 2.24[2.04-2.47]<br>P<10<sup>-5</sup> |
| High PRS vs low PRS within strata of disability | 1.10[1.04-1.17]<br>P=0.023 | 1.36[1.23-1.49]<br>P<10<sup>-5</sup> |  |
| Ms | 1.23, [1.10-1.38], P=3.3X10<sup>-4</sup>. |  |  |
| RERI | 0.55, [delta:0.32-0.77], [mover:0.33-0.78], P<10<sup>-5</sup> |  |  |
| SI | 0.22, [delta:0.14-0.30], [mover:0.14-0.29], P<10<sup>-5</sup> |  |  |
| AP | 1.59, [delta:1.32-1.91], [mover:1.32-1.91], P<10<sup>-5</sup> |  |  |

Note: Disability: other serious medical condition disability diagnosed by a doctor, Multiplicative scale, RERI= relative excess risk due to interaction, AP= attributable proportion, and SI= synergy index.

**Table 2.**
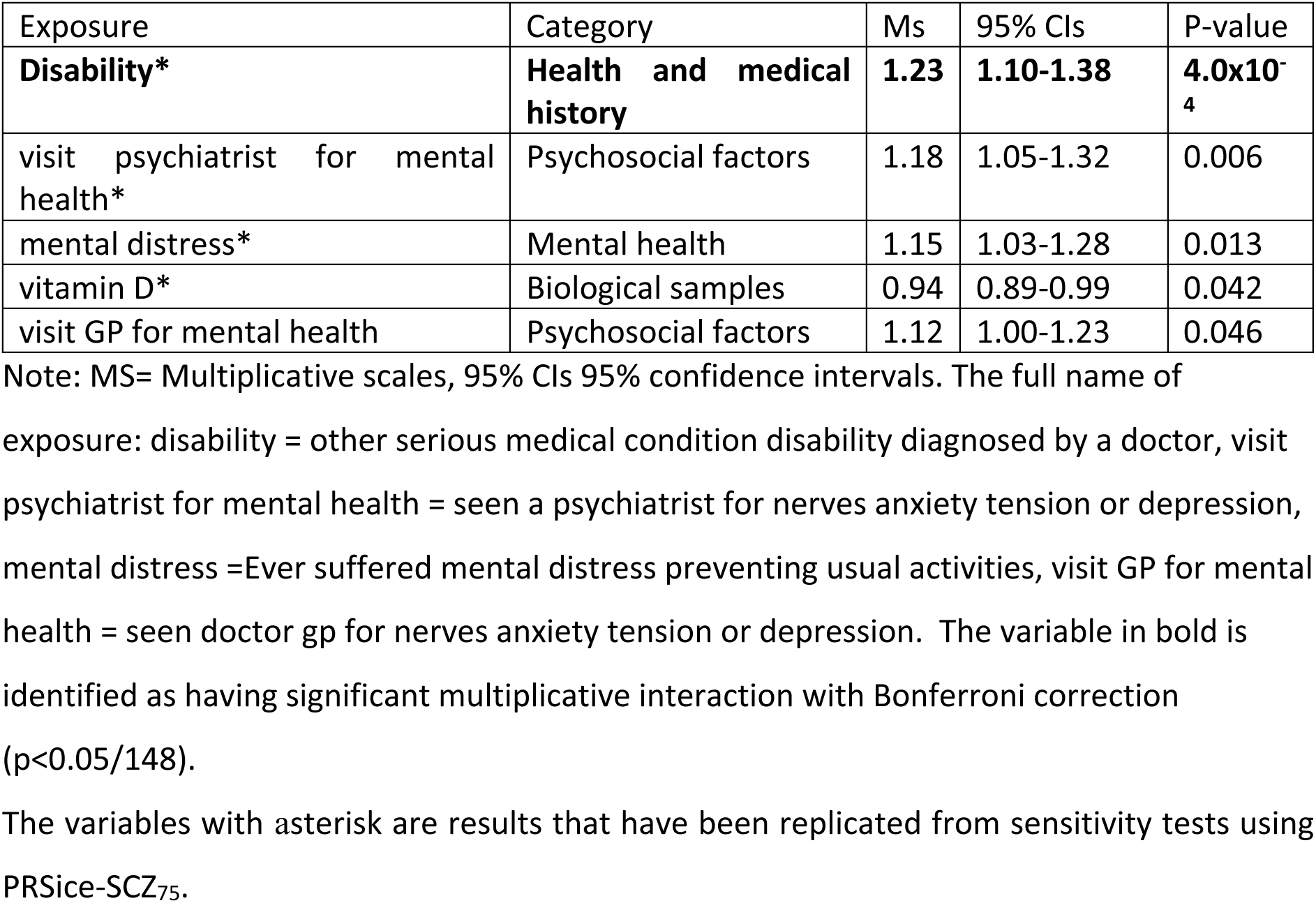
Significant multiplicative interactions of PRS-cs-auto-SCZ_75_ and exposures on psychotic experiences were identified using an exposome-wide interaction study.

| Exposure | Category | Ms | 95% CIs | P-value |
| --- | --- | --- | --- | --- |
| **Disability\*** | **Health and medical history** | **1.23** | **1.10-1.38** | **4.0x10<sup>-4</sup>** |
| visit psychiatrist for mental health* | Psychosocial factors | 1.18 | 1.05-1.32 | 0.006 |
| mental distress* | Mental health | 1.15 | 1.03-1.28 | 0.013 |
| vitamin D* | Biological samples | 0.94 | 0.89-0.99 | 0.042 |
| visit GP for mental health | Psychosocial factors | 1.12 | 1.00-1.23 | 0.046 |

Note: MS= Multiplicative scales, 95% CIs 95% confidence intervals. The full name of exposure: disability = other serious medical condition disability diagnosed by a doctor, visit psychiatrist for mental health = seen a psychiatrist for nerves anxiety tension or depression, mental distress =Ever suffered mental distress preventing usual activities, visit GP for mental health = seen doctor gp for nerves anxiety tension or depression. The variable in bold is identified as having significant multiplicative interaction with Bonferroni correction ( $p < 0.05/148$ ).
The variables with asterisk are results that have been replicated from sensitivity tests using PRSice-SCZ<sub>75</sub>.

### Additive interaction

Among the 148 variables, significant additive interactions were found for seven exposures (disability, mental distress, sadness, help for mental distress, sleeping difficulties, visiting a GP for mental health and visiting a psychiatrist for mental health) (**Figure 1 & Table 3**). Similar to the multiplicative interaction analyses, disability interacted with PRS-cs-auto-SCZ_75_ on an additive scale (**eTable 1**: RERI,0.55; 95%CI, 0.32-0.77; SI, 0.22; 95%CI, 0.14-0.30; AP, 1.59; 95%CI, 1.30-1.91; all P < 0.05/148). The MOVER method identified similar confidence intervals (**eTable 5**).

**Figure 1.**
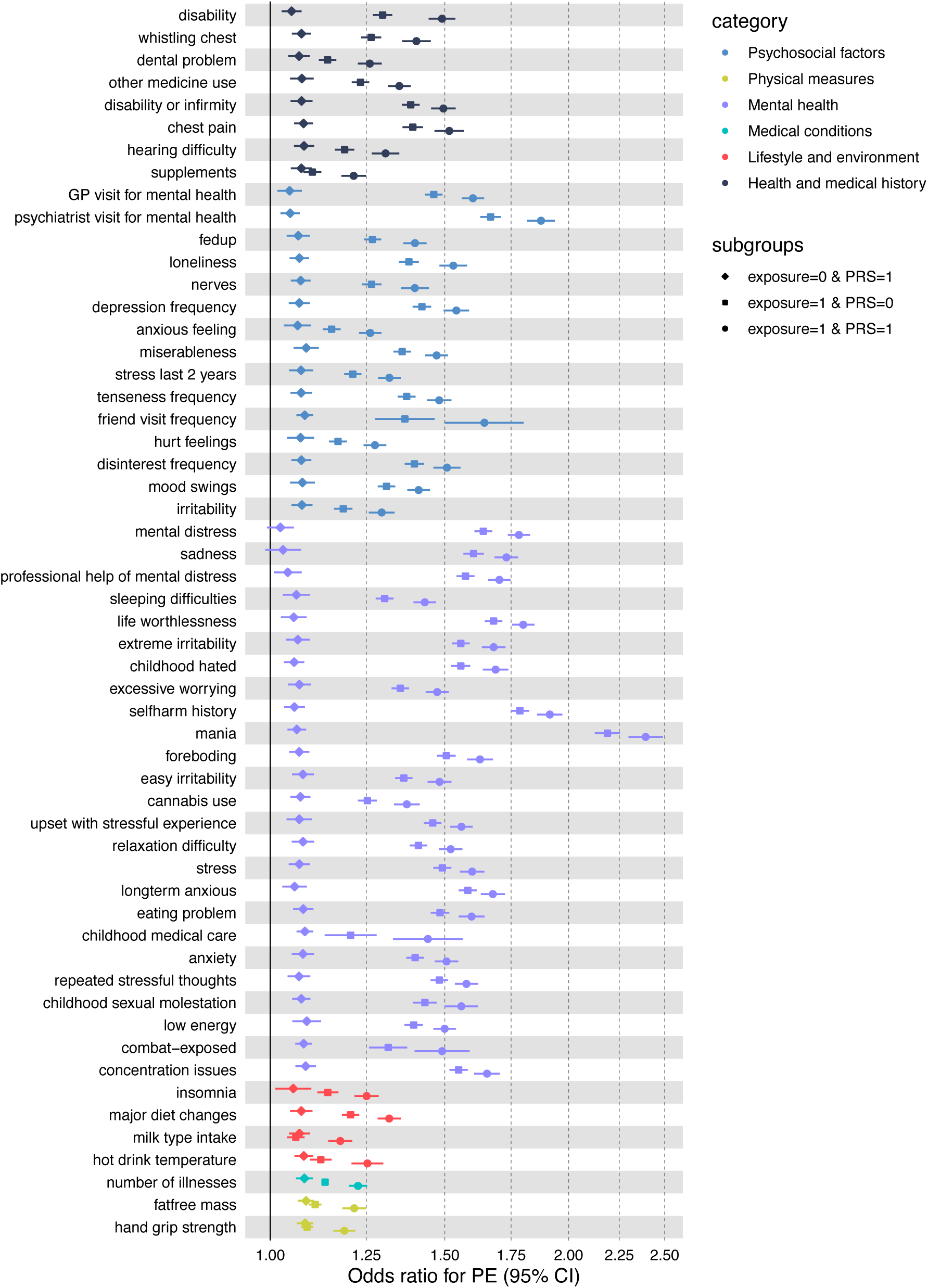
Odds ratios of PE in 55 exposures and PRS subgroups. 55 exposures are nominal significant from the additive interaction test.

**Table 3.**
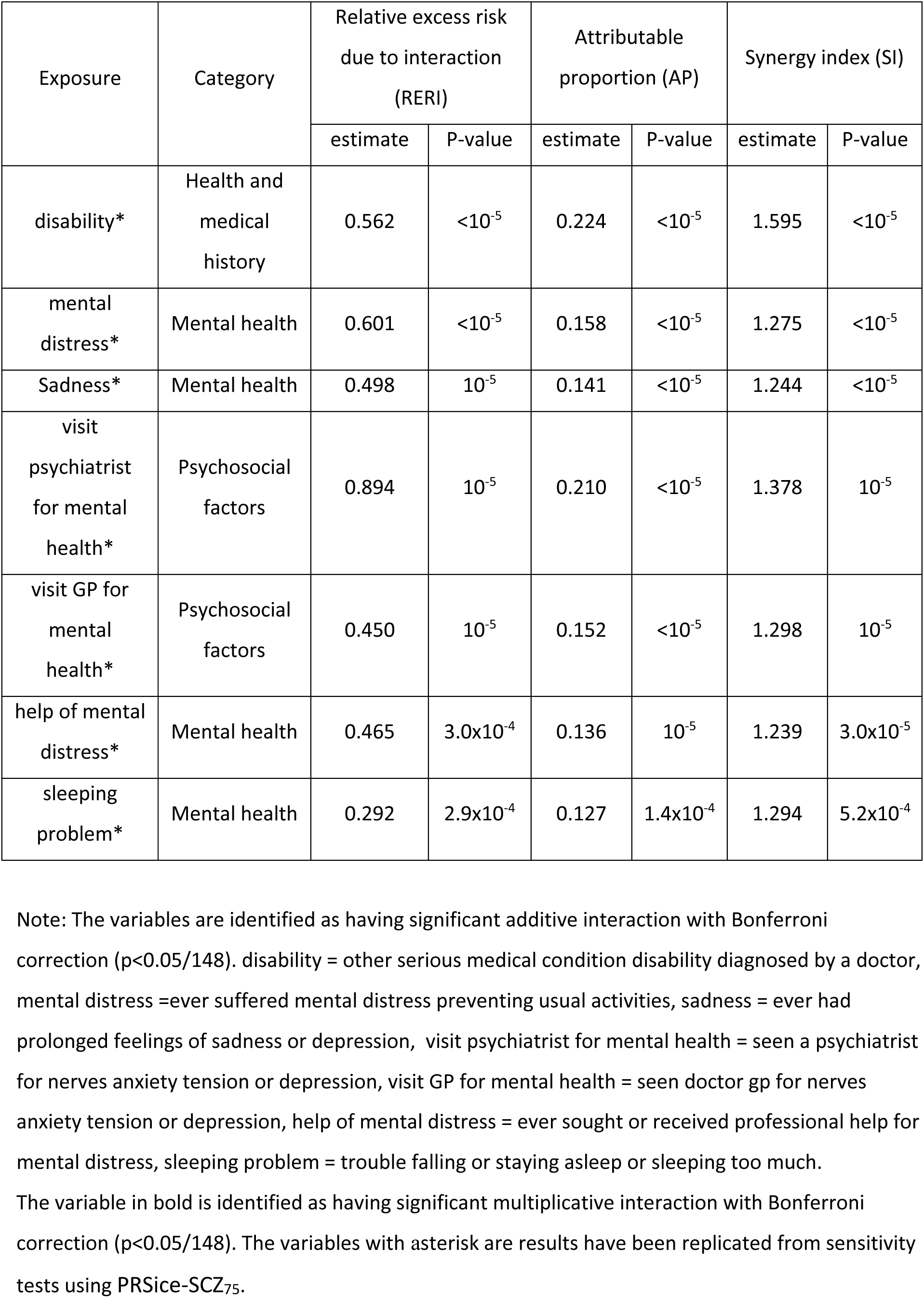
Significant additive interaction between PRS-cs-auto-SCZ_75_ and exposures on psychotic experiences were identified using an exposome-wide interaction study.

An additional 48 interactions were detected with nominal significance levels (**eTable 5 and eFigure 3**). The majority of these exposures were from the UKB mental health (n=25) and psychosocial factors (n=15) categories, including cannabis use, self-harm, eating problems, sexually molested as a child, and loneliness isolation. Furthermore, interactions with exposures from the following categories were found: health and medical history (n=8: e.g. chest, dental, infirmity, hearing problem and vitamin supplements), lifestyle and environment (n=4: insomnia, diet change, milk type used, and hot drink temperature), physical measures (n=2: fat mass and hand grip strength) and medical conditions (number of illness).

The sensitivity analyses using PRSice-SCZ_75_ confirmed the seven significant additive interactions. Furthermore, 39 out of the 48 nominal significant additive interactions were confirmed (**eTable 5**).

### Gene-environment correlations

The correlation analyses found small (r^2^ range from −0.021 to 0.058) but significant (P<2.02×10^-4^) correlations between 102 exposures and PRS-cs-auto-SCZ_75_ (**eTable 6**). Ninety-four (r^2^ range from −0.028 to 0.042) of these correlations remained significant using PRSice-SCZ_75_ in sensitivity tests.

Focusing on the exposures with significant interactions with PRS, disability and sleeping problems were not correlated with PRS-SCZ. Although the rest of the exposures that interacted with PRS-SCZ were positively correlated with PRS-cs-auto-SCZ_75_, the magnitude of the correlations was very small (<0.04). These correlations were replicated in the sensitivity tests using PRSice-SCZ_75_.

### GxE interaction with birth weight and smoking

Birth weight was initially excluded from the previous XWAS due to a missing rate of >10%. Smoking status, pack years of smoking and maternal smoking around birth were also excluded due to collinearity, missingness, and being a follow-up variable, respectively. However, we extracted these variables to replicate previous findings and estimated the additive and multiplicative interactions with PRSice-SCZ_75_ on PE (**eTable 7 and eFigure 4**). Among these four variables, only a nominally significant additive interaction of smoking status with PRS-SCZ on PE was found (RERI,0.13; 95%CI, 0.014-0.266; P=0.038).

## Discussion

To the best of our knowledge, this study represents the most extensive systematic inquiry into the exposome-wide gene-environment interaction of PE. It encompasses several sequential analytical steps, including an exposome-wide association study conditional on PRS-SCZ, an exposome-wide gene-environment interaction investigation, an exposome-wide gene-environment correlation estimation, and replication of previous GxE analyses.

Our exposome-wide gene-environment interaction study identified significant multiplicative and additive interactions between disability and genetic risk of schizophrenia on PE, as well as six significant additive interactions: help and treatment-seeking behaviours, mental distress and sleep problems. Besides the significant interactions, four multiplicative and 48 additive nominally statistically significant interactions were identified, mainly in the domains of physical health outcomes, non-psychotic disorders, mental distress, stress, trauma, help and treatment-seeking behaviors, and sleep problems. Overall, more significant additive interactions were detected compared to multiplicative interactions. Compared to multiplicative interaction tests, additive interaction tests may offer greater statistical power and reveal more interpretable results from biomedical and epidemiological data (41)

Our study found that the impact of physical disability on PE increased with higher PRS-SCZ, as revealed by both multiplicative and additive GxE models. To the best of our knowledge, this is the first report indicating that the sensitivity to adverse physical conditions is moderated by PRS-SCZ. PE, an indicator of general health, has been associated with increased risk for disability across a broad range of functional domains including social-, role-, cognitive functioning, mobility, and self-care (42). We showed that both conditional XWAS tests and XWIS models, which include PRS-SCZ and GxE, explained more variance of PE than models testing only environmental factors. This finding supports the idea that polygenic risk, poor physical health, and their combined influence are associated with subthreshold psychosis expression.

Additionally, we identified nominally significant additive interactions with milder physical health issues like chronic illness or recent fatigue, with smaller RERI (0.23, 0.17) and AP (0.09, 0.07) values compared to Bonferroni significant interaction of disability with more serious condition (RERI=0.56, AP=0.24), indicating stronger GxE effects with severe health outcomes. Furthermore, gene-environment interactions have also been detected for other physical health outcomes, such as wheeze or whistling in the chest in the last year, chest pain, dental problems, taking other prescription medications, number of self-reported noncancer illnesses, and hearing problems. Our findings highlight the gene-environment interaction of serious medical conditions or disabilities with genetic propensities for schizophrenia on PE. This supports a conceptual framework where underlying (nonspecific) immune dysfunction (e.g. autoantibodies, T- and B cells), with an estimated heritability of 30% (43), might serve as a foundational mechanism leading to a broad spectrum of health outcomes, including psychosis, contingent on disease burden. Of note, the gene-environment interactions became stronger with increasing severity of physical condition, suggesting a dose-response relationship where increased disease burden might exacerbate PE, akin to sickness behavior during illness (44, 45). In this regard, immune system dysregulation and neuroinflammation might be the culprit for behavioural and functional impairments (46).

In the XWIS study, three significant additive interactions were identified for treatment-seeking behavior linked to mental health problems: seeing a psychiatrist for nerves, anxiety, tension or depression, seen doctor/gp for nerves, anxiety, tension or depression, and ever sought or received professional help for mental distress. In accordance with our findings, this suggests that targeting high PRS-SCZ and help-seeking individuals may aid in intervening in psychotic disorders. Furthermore, our findings identified suggestive interactions of well-known exposures such as cannabis use, self-harm, medical prescription, and sexually molested as a child, which is consistent with previous studies with independent samples (13, 30) Additionally, suggestive interactions were identified for

According to the diathesis-stress theory, it is crucial to identify cumulative stressors that contribute to the manifestation of psychiatric symptoms in vulnerable populations like those with PE. Illness behaviour, characterized by patterns of seeking professional help (47), and diathesis (vulnerabilities), both play pivotal roles in understanding why certain individuals are more prone to develop psychiatric symptoms. Individuals with a high genetic predisposition to psychosis may exhibit intrinsic issues with information processing and misinterpretations, potentially intensifying their response to environmental stressors and increasing the likelihood of experiencing psychiatric symptoms and seeking professional help. Cumulative stressors, such as ongoing life difficulties and acute stress events, can exacerbate vulnerability, potentially triggering adverse illness behaviors and increasing the need for intervention. It is critical to identify and manage cumulative stressors in genetically vulnerable populations, particularly those with high genetic liability for schizophrenia. Overall, our results align with the diathesis-stress model theory, suggesting that a combination of genetic predisposition and environmental stress contribute to the vulnerability of PEs.

Several prior investigations have evaluated the interplay between PRS-SCZ and environmental variables underlying PE. However, these studies have predominantly focused on a limited number of environmental factors such as stress (48), smoking behavior (27), and birth weight, which have not been verified in independent cohorts(26). In our study, we have replicated previous findings with suggestive interaction for stress (“felt very upset when reminded of stressful experience in past month and avoided activities” or “situations because of previous stressful experience in past month”) and smoking status.

Previous exposome-wide analyses identified 148 exposures associated with PE. The subsequent conditional cross-phenotype-wide association study (XWAS) reaffirms that the relative impact of genetic factors on PE (with only 0.2% variance explained by PRS-SCZ) is notably lower compared to environmental exposures. Our results were consistent with the findings of a twin study suggesting that environmental factors might play a greater role than genetic factors in the etiology of PE (20). Conversely, even after adjusting for exposures associated with PE, a significant association of PRS-SCZ with PE persisted, underscoring the importance of genetic predispositions to schizophrenia on PE.

Our research has several strengths. First, the UK Biobank’s deep phenotyping and large sample size provide the requisite statistical robustness to discern subtle GxE, even within complex multifactorial outcomes such as PE. This capability enables the identification of interactions with heightened precision. Second, we employed two widely recognized methods for PRS calculation: PRS-cs-auto and PRSice2. The PRS-cs-auto generation method allows for the efficient processing of vast amounts of genetic data and yields more statistically robust results, particularly in the context of larger sample sizes. Additionally, we utilized PRSice2 to generate PRS-SCZ, employing a liberal p-value threshold of 0.05 for sensitivity analyses, thereby enhancing the predictive power of genetic scores. Third, our study benefited from access to the most extensive GWAS summary statistics available to date. The variance of PE explained by PRS-SCZ in our study (0.2%) is larger than another study using summary statistics from Psychiatric Genomic Consortium freeze 2 (PGC2) (27). Although our systematic approach was designed to mitigate biases and increase reproducibility, it was not without limitations. First, the sequential replication procedure and stringent multiple-testing correction might have inadvertently increased the likelihood of type II errors. Conversely, statistically significant yet trivial effects can also emerge in analyses of large datasets. Second, we have not investigated any subtypes of PE; therefore, the contribution of genetic risk and exposures on specific types of PE remain unknown. Last, the proportion of variance of PE explained by PRS-SCZ was minimal (<2%). Additional investigation is necessary to clarify the other genetic contributors (rare variants and Copy Number Variants) to phenotypic variance. Future research may utilize deep sequencing and prospective designs to generate more robust evidence.

## Conclusion

The current study marks the first documentation of numerous exposures associated with psychotic experiences, after adjusting for polygenic risk for schizophrenia. These findings reveal preliminary evidence for gene-environment interaction in psychotic experiences and suggest that genetic vulnerability and exposures might represent intertwined pathways leading to psychosis. Our findings support the diathesis-stress theory, and underscore the necessity of evaluating both environmental and genetic influences in conjunction to elucidate biological mechanisms underlying psychosis.

## Data availability

All results data generated and analyzed during this study are included in the supplementary materials accompanying this manuscript. These supplementary materials provide the complete dataset necessary to interpret, verify, and extend the research presented in the article.

For any additional information or access to specific datasets beyond what is provided in the supplementary materials, reasonable requests can be made to the corresponding author.

## Acknowledgments

Drs Lin and Guloksuz had full access to all of the data in the study and take responsibility for the integrity of the data and the accuracy of the data analysis. Concept and design: Drs Lin and Guloksuz. Acquisition, analysis, or interpretation of data: All authors. Drafting of the manuscript: Dr Lin. Statistical analysis: Dr Lin. Critical revision of the manuscript for important intellectual content: Lin, Pries, Arias-Magnasco, Klingenberg, Guloksuz. Obtained funding: Rutten, Guloksuz. Supervision: Dr Guloksuz.

