## Supplementary material for "Exposome-wide gene-environment interaction study of psychotic experiences in the UK Biobank"

**Supplementary materials**

**Quality control and pre-processing of the dataset**

UK Biobank (UKB) is a large prospective database containing genetic, lifestyle, and health-related information from half a million UK participants. The UKB database is globally accessible to researchers who are undertaking health-related research that is in the public interest. UKB recruited 500,000 people aged 40-69 years in 2006-2010 from across the UK. With their consent, participants provided lifestyle and health information as well as blood, urine, and saliva samples, which were collected and stored for future analysis. The UKB resource was opened for research use in April 2012. UKB is supported by its founding funders the Welcome Trust and UK Medical Research Council, as well as the British Heart Foundation, Cancer Research UK, Department of Health, Northwest Regional Development Agency, and Scottish Government.

The current study (UKB project number: 55392) analyzed participants with complete data on the mental health questionnaire that assessed psychotic experiences (n=155,247; 57% female; mean age=55.94 and standard deviation [SD] = 7.74 years). The following procedure was applied to prepare the dataset for the systematic analyses of the non-genetic correlates of psychotic experiences (PE). Initially, the UKB dataset included 23,089 variables. We first excluded 15,800 variables based on the following reasons using the information provided in the UKB showcase (see eTable 1 for details): repeated measurements after the first array of variables with multiple data items were excluded (“Comes after first array”: n=7,684); variables’ value type were “Compound” (n=30), “Date” (n=1,709), or “Time” (n=27); only reported by (specific to) female participants (“Female only”, n= 190); “Follow-up” (branch) queries (n=2,603); “Genetic” (n=9); only reported by (specific to) male participants (“Male only”; n=38); variables’ item type were “Bulk” (n=94) or “Records” (n=10); variables’ strata type were “Auxiliary” (n=2,806) or “Supporting” (n=600). We further excluded variables that showed no variance (“No variance”; n=371). All above-mentioned excluded variables (n=16,171) were listed in eTable 1. The remaining 6,918 variables contained several instances of the same variable. We used information from the first instance when available. If values in the first instance were missing, these were replaced with follow up instances when they were available. After the pre-processing1, the initial raw dataset included 4,678 independent variables. Then, we excluded variables that had missing rates above a priori set missing rate cutoff > 0. 1. 303 variables remained. All non-ordered categorical variables were dichotomized with the most frequent category denoted by “0” and the rest by “1” (e.g. “attendance/disability/mobility allowance” was coded with “attendance allowance” = 1, “Disability living allowance” = 1, “Blue badge” = 1, “None of the above” = 0, “Prefer not to answer” = NA, “Do not know” = NA). To avoid potential sparsity and guided by a previous study1, numeric variables with <10 values were dichotomized with the lowest value denoted by “0” and the rest by “1”. The numeric variables with ≥10 values were treated as continuous to avoid loss of possible meaningful information and transformed into z-scores. Following this, we checked for collinearity and excluded one of two variables from a highly-correlated pair (r2 >0.9) and retained the variables that were less frequently highly-correlated with other variables in the dataset by using the R program: findCorrelation (from the caret package4). We excluded 56 highly-correlated variables. Eventually, the final number of variables that were included in the exposome-wide analyses was 247.

**PRS-cs-auto**

PRS -cs-auto was generated by applying a Bayesian framework method that uses continuous shrinkage (cs) on SNP effect sizes. PRS-cs-auto can accommodate diverse underlying genetic architectures because the continuous shrinkage priors allow for marker-specific adaptive shrinkage. In addition, PRS-cs-auto can accurately model local LD structures and provide substantial computational improvements by conjugating block update of the SNP effect sizes in posterior inference. Furthermore, PRS-cs-auto is built to scale with the size of the dataset. As the sample size increases, the algorithm can efficiently process the additional data without a significant increase in computational time or resource requirements. This scalability enables us to analyze larger sample sizes and obtain more statistically robust results. PRS-cs-auto is robust to varying genetic architectures, provides substantial computational advantages, and enables multivariate modelling of local linkage disequilibrium patterns.

**PRSice**

First, all overlapping SNPs between GWAS summary statistics (base dataset, 1000 Genomes phase I reference dataset: <https://www.internationalgenome.org/data/>) And our target dataset was selected. Then the following SNPs were excluded using PLINK2 (49): 1) insertion or deletion, ambiguous SNPs; 2) SNPs with minor allele frequency (MAF) <0·01 and SNPs with imputation quality (R2) <0·8 in both training dataset and target datasets; 3) SNPs located in complex-LD regions (**Supplemental Table S1**) (50). These SNPs were clumped in two rounds; round 1 with the default parameters (physical distance threshold 250kb and LD threshold (R2) <0·5; round 2 with a physical distance threshold of 5,000kb and LD threshold (R2) <0·2. Odds ratios in the summary statistics were log-converted to beta values. PRS was calculated using PRSice2 for all SNPs with p-value threshold at 0∙05 (51).

**Additive and multiplicative interaction**

In this study, interaction effects were assessed on different scales, specifically additive and multiplicative scales, with the choice of scale playing a critical role in interpreting the combined effects of variables. While measures of multiplicative interaction, which involve multiplying the individual effects of variables, are commonly reported in practice for convenience, their selection is often made without careful consideration of preference (52). Despite the additional effort required to obtain measures of additive interaction within current software packages, the preference for additive interaction tests arises from their ease of interpretation and communication compared to multiplicative interactions.

The detection of additive interaction, where the combined effects of variables are summed, can be instrumental in identifying subgroups that derive the greatest benefits from interventions. This information facilitates targeted and efficient allocation of healthcare resources. Notably, additive interaction tests may exhibit greater power in detecting specific types of interactions or effects, as evidenced in this study where more significant additive interactions were detected compared with multiplicative interactions. Moreover, additive interactions are highlighted for their potential robustness and reliability across various study designs and data structures, contributing to the observation of more substantial findings in the current study. By providing insights into the absolute differences in outcomes between subgroups, additive interaction proves invaluable in assessing the practical impact of interventions and elucidating the underlying mechanisms of interactions.

**Supplemental Table S1. 20 complex-LD (linkage disequilibrium) regions and long-range LD regions that were excluded from Polygenic Risk Score-analysis (50).**

| Chromosome | Base pair position (start point to end point) |
| --- | --- |
| 1 | 48000000-52000000 |
| 2 | 86000000-100500000 |
| 2 | 183000000-190000000 |
| 3 | 47500000-50000000 |
| 3 | 83500000-87000000 |
| 5 | 44500000-50500000 |
| 5 | 129000000-132000000 |
| 6 | 25500000-33500000 |
| 6 | 57000000-64000000 |
| 6 | 140000000-142500000 |
| 7 | 55000000-66000000 |
| 8 | 8000000-12000000 |
| 8 | 43000000-50000000 |
| 8 | 112000000-115000000 |
| 10 | 37000000-43000000 |
| 11 | 87500000-90500000 |
| 12 | 33000000-40000000 |
| 20 | 32000000-34500000 |
| 8 | 8135000-12000000 |
| 17 | 40900000-45000000 |

**eFigure 1. Schematic Overview of the Study Design.**

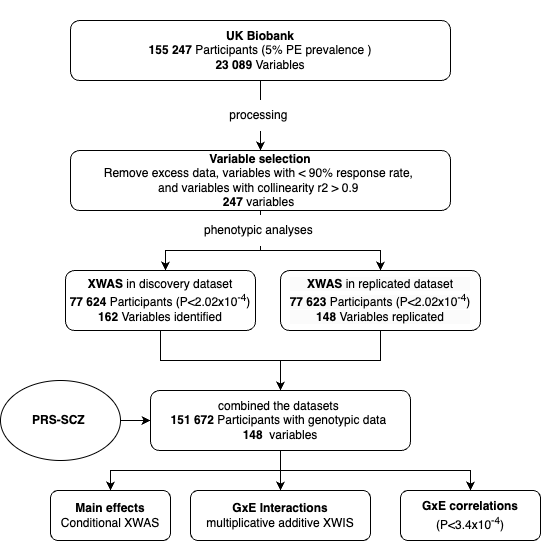

**eFigure 2.** The Odds ratios (ORs) of 148 exposures on PE in the previous XWAS models without PRS correction compared to the odds ratios of 148 exposures on PE in the PRS-cs-auto-SCZ_75_ conditional XWAS models.

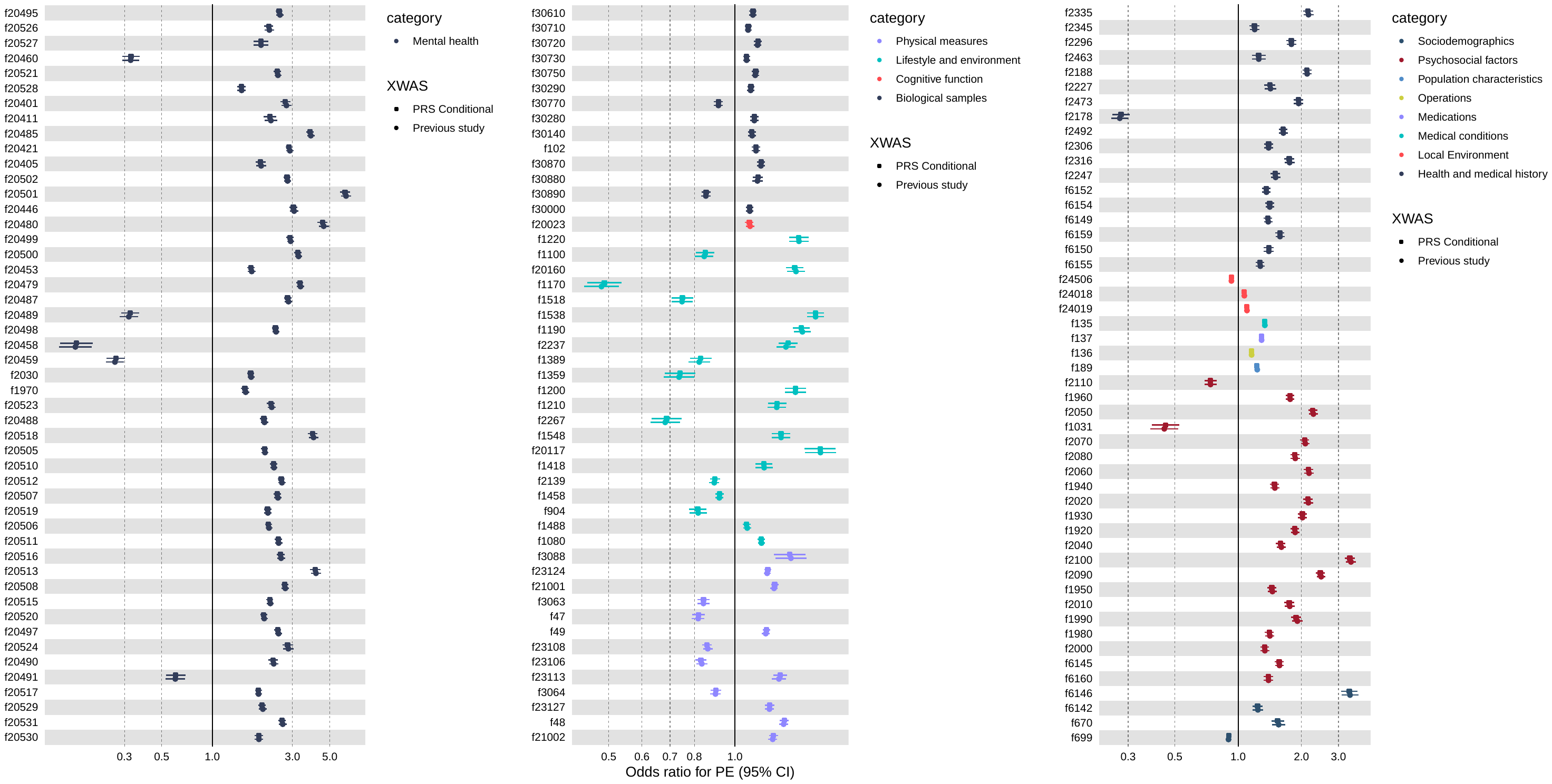
Note: The round dots represent ORs of exposures on PE without adjusting PRS-SCZ from the previous XWAS study. The diamonds represent ORs of exposures on PE PRS-SCZ as covariates from PRS conditional XWAS study.

**eFigure 3.** The associations between 48 exposures with PRS-cs-auto-SCZ_75_ on the risk of PE with nomial Additive Interaction (P<0.05).
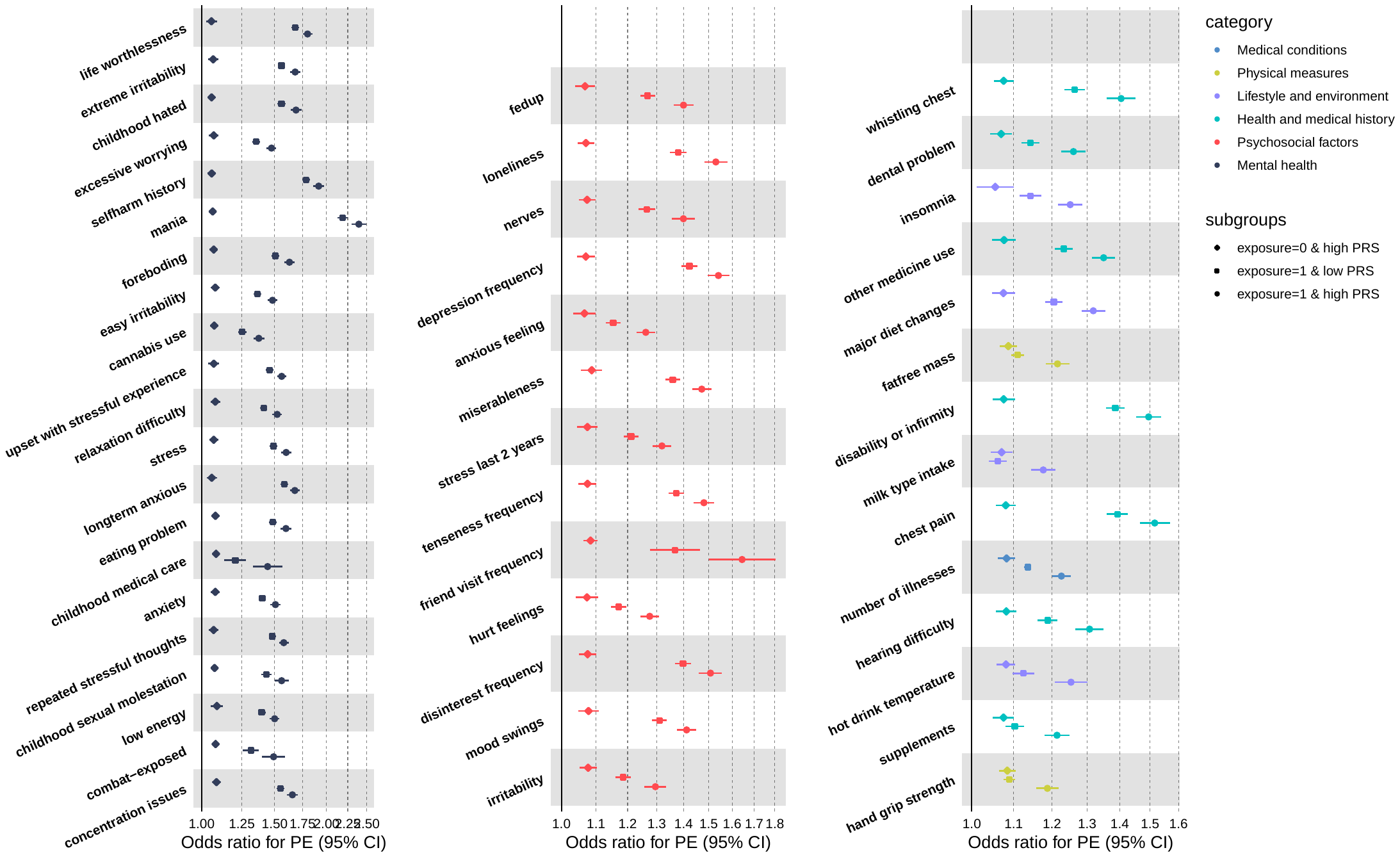

**eFigure 4.** Associations between birth weight and smoking behaviour with PRS-cs-auto-SCZ_75_ on risk of PE.

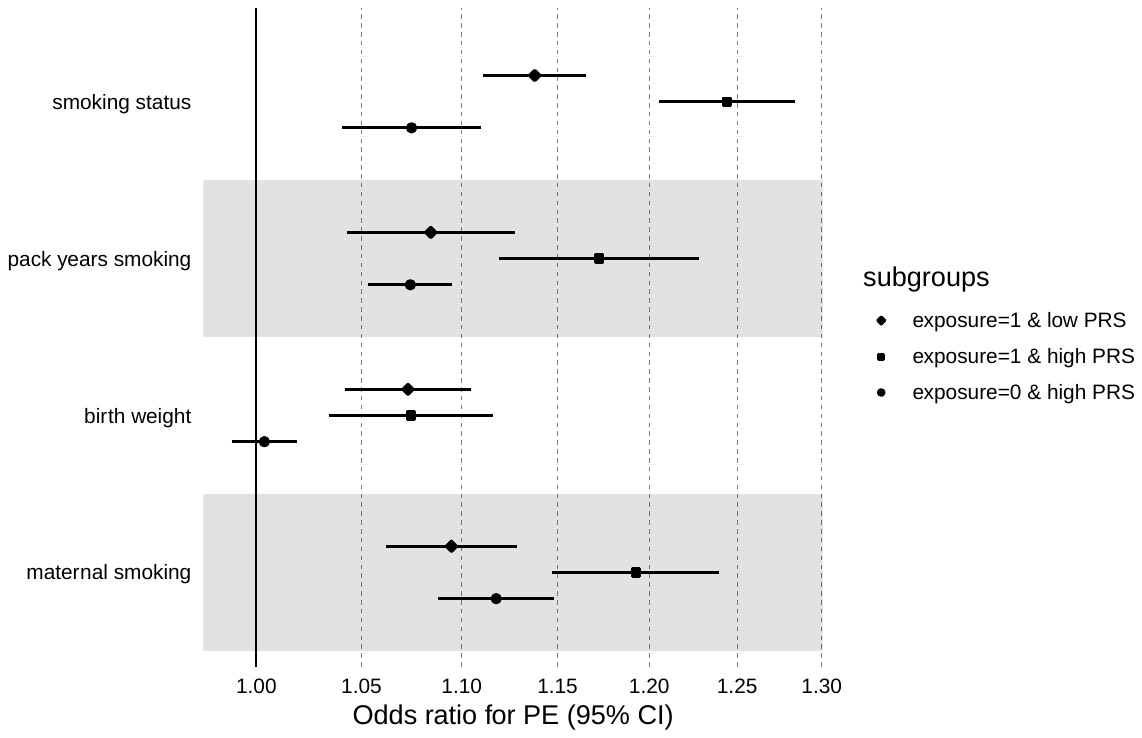
